# Preoperative Exposure to Fine Particulate Matter and Risk of Postoperative Complications: A Single Center Observational Cohort Bayesian Analysis

**DOI:** 10.1101/2024.08.13.24311943

**Authors:** John F. Pearson, Cameron K. Jacobson, Calvin S. Riss, Matthew J. Strickland, Longyin Lee, Neng Wan, Tabitha M. Benney, Nathan L. Pace, Ben K. Goodrich, Jonah S. Gabry, John V. Pham, Cade K. Kartchner, Jake S. Wood, Michael H. Andreae

## Abstract

**Background:** While exposure to fine particulate matter air pollution (PM_2.5_) is known to cause adverse health effects, its impact on postoperative outcomes in US adults remains understudied. Perioperative exposure to PM_2.5_ may induce inflammation that interacts insidiously with the surgical stress response, leading to higher postoperative complications.

**Methods:** We conducted a single center, retrospective cohort study using data from 49,615 surgical patients living along Utah’s Wasatch Front and who underwent elective surgical procedures at a single academic medical center from 2016-2018. Patients’ addresses were geocoded and linked to daily Census-tract level PM_2.5_ estimates. We hypothesized that elevated PM_2.5_ concentrations in the week prior to surgery would be associated with an increase in a bundle of major postoperative complications. A hierarchical Bayesians regression model was fit adjusting for age, sex, season, neighborhood disadvantage, and the Elixhauser index of comorbidities.

**Results:** Postoperative complications increased in a dose-dependent manner with higher concentrations of PM_2.5_ exposure, with a relative increase of 8% in the odds of complications (OR=1.082) for every 10ug/m^3^ increase in the highest single-day 24-hr PM_2.5_ exposure during the 7 days prior to surgery. For a 30 fold increase in PM_2.5_ (1 ug/m^3^ to 30ug/m^3^) the odds of complication rose to over 27% (95%CI: 4%-55%). The association persisted after controlling for comorbidities and confounders; our inferences were robust to modeling choices and sensitivity analysis.

**Conclusions:** In this large Utah cohort, exposure to elevated PM_2.5_ concentrations in the week before surgery was associated with a dose-dependent increase in postoperative complications, suggesting a potential impact of air pollution on surgical outcomes. These findings merit replication in larger datasets to identify populations at risk and define the interaction and impact of different pollutants. PM_2.5_ exposure is a potential perioperative risk factor and, given the unmitigated air pollution in urban areas, a global health concern.

## Introduction

Short-term exposure to air pollution, especially fine particulate matter of less than 2.5 microns in size (PM_2.5_) is a well-established risk factor for cardiovascular, respiratory, metabolic, and neurologic, morbidity, and mortality worldwide.^1–6^ The increasing prevalence of wildfires induced by climate change worsens exposure to fine particulate matter in the US,^7^ while millions globally continue to suffer under high levels of PM_2.5_^8,9^. There is emerging evidence that air pollution and the stress response to surgery (**SRS**) interact insidiously as there is overlap between the SRS and the oxidative stress-induced injuries of air pollution. SRS results in pulmonary trauma, hemodynamic stress, and proinflammatory cytokines,^10–16^ while air pollution similarly results in increased risk of thrombosis, airway inflammation, and alterations in the immune response.^17,18^

The mechanism by which air pollution induces systemic inflammation is well established: small airborne pollutants enter through the lungs or skin^19^ and disseminate across the body. Such pollutants can be found in every organ system, including the lungs, brain, the heart, and gastrointestinal tract among others^20^. The impact of individual patient air pollution exposure (IPAPE) thus may lead to an inflammatory response that compounds the SRS^13,21^, including pulmonary inflammation, endothelial dysfunction, thrombosis, and membranous nephropathy (Appendix Figure 1).^10,11,15,16^ Only a small number of studies^22–26^ examined the impact of IPAPE on perioperative outcomes and those that did were limited to specialized populations (Appendix Table 1: Rigor of Prior Research). A recent analysis of over 1.3 million patients in China found increased 30-day postoperative mortality was associated with higher preoperative PM_2.5_ concentrations at the city-level on day of surgery, especially among patients with preexisting cardiopulmonary conditions^23^. Yet no study examined major post-operative medical complications in any U.S. adult surgical population.

The Salt Lake metropolitan region of Northern Utah is an ideal setting to study acute pollution exposures. Due to the various air pollution and weather events common to the region (e.g. inversion, wildfires) which quickly clear when the weather changes, the random exposure of patients to variable air pollution in the days prior to surgery leads to a quasi-natural experiment as patients are scheduled for surgery regardless of air pollution exposure (Figure 1: The rapidly changing pollution levels along the Wasatch Front).

**Figure 1:**
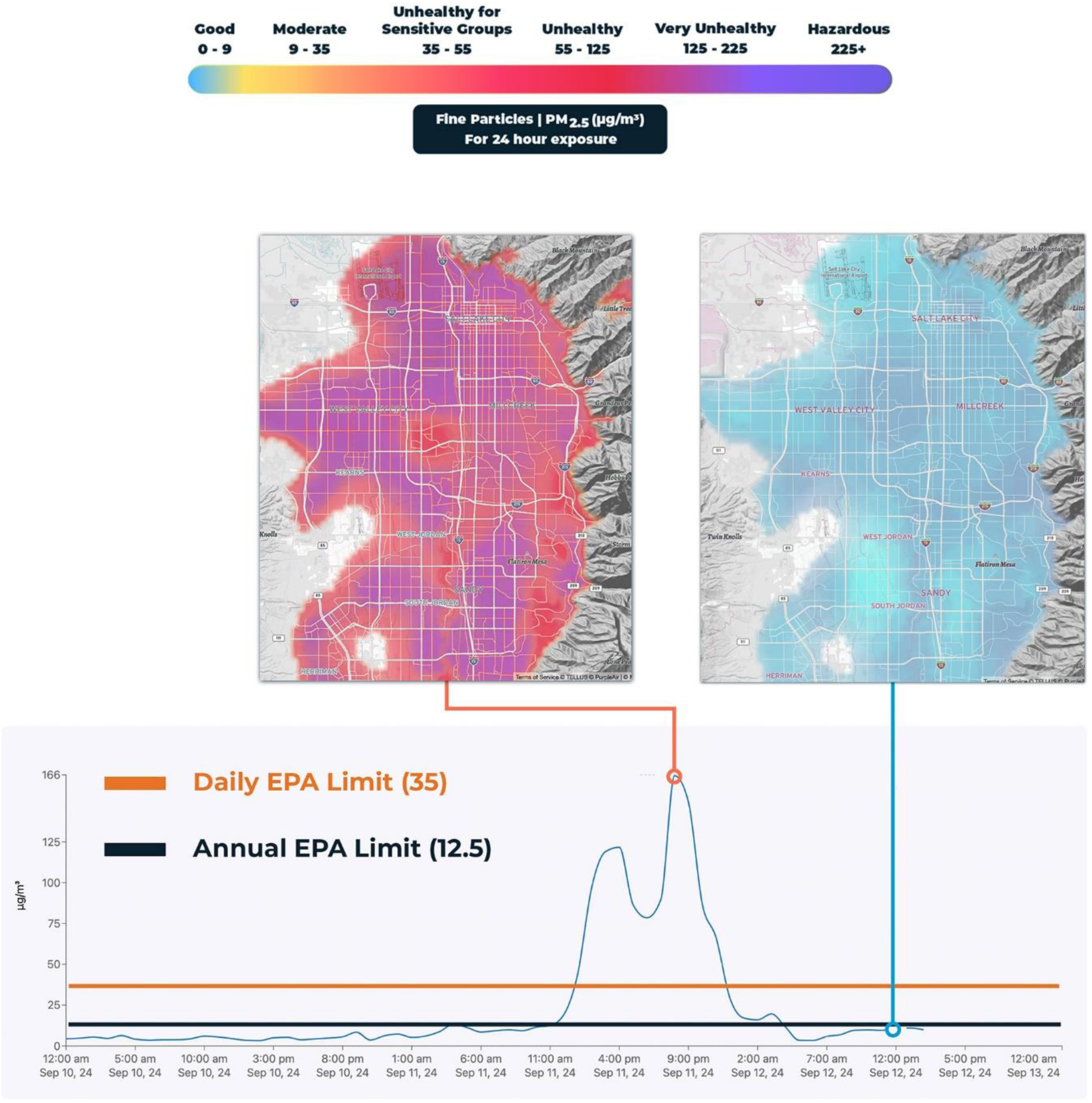
Rapidly changing pollution levels along the Wasatch front. Figure 1 illustrates the rapidly changing pollution levels along the “Wasatch Front” mountain range in Salt Lake City, UT. The unique mountain geography of the “Wasatch Front,” along with five oil refineries, multiple industrial facilities, 2.2 million people, the intersection of two major interstate highways, and a national train depot, work together to produce the increased occurrence of extreme pollution events in the region, which dramatically impact ambient PM_2.5_ concentrations^32,58,59^. This results in inversion conditions during the winter, with warm air aloft the valley trapping cold air and pollutants in the densely populated valley below. Similarly, in the summer when wildfires throughout the American West dominate pollution exposures, the geological bowl produced by the intersection of multiple mountain ranges that comprise the Wasatch Front act as a shield that accumulates wildfire smoke in the metropolitan region. In both instances, low pressure systems tend to rapidly clear the area of pollutants, at random. As a result, patients are quasi-randomly exposed to high or low pollution levels, while their surgery scheduling is disregarding this exposure, leading to a natural experiment. The left map illustrates high observed pollution levels (red/purple for unhealthy high pm 2.5 levels) due to a recent wildfire event on September 11-12, 2024 while the right map demonstrates how a change in weather patterns cleared the air, leading to suddenly much lower pollution exposures (blue). The graph demonstrates this timeline of rapidly changing 2.5 small particle pollution levels, with markers for the dates of the maps.

Our objective was to estimate the impact of preoperative particulate matter (PM_2.5_) air pollution on a bundle of post-operative complications in a local cohort of cases in the national perioperative electronic health registry MPOG.org. We hypothesized that exposure to PM_2.5_ in the 7-day period prior to surgery results in an increased risk of a bundle of post-operative complications, after controlling for comorbidities, confounders, and other well documented drivers of post-operative complications^27,28^ in a hierarchical Bayesian regression model.

## Materials and Methods

### Study Design and Data Sources

We performed a single center cohort analysis of the University of Utah local Multi-Center Perioperative Outcomes Group (MPOG)^29^ electronic health registry, which was supplemented with data from the University of Utah Health’s Epic database for the period January 1, 2016 to December 31, 2018. We adhered to the Strengthening the Reporting of Observational Studies (STROBE) statement and principles^30^. The study was approved as Exemption Category 4 by the Institutional Review Board at the University of Utah, with IRB approval number 00142167. Prior to access of data, we wrote of our intention to analyze the influence of environmental factors and air pollution on postoperative outcomes with the University of Utah IRB. During the preparation of this work the authors used OpenAI’s ChatGPT for assistance with writing clarity and literature search. After using this service, the authors reviewed and edited the content and take full responsibility for the content of this publication.

### Study Population: Inclusion and Exclusion Criteria

Extracting information from our Epic and local MPOG instances we included elective and non-emergent general anesthesia cases performed at University of Utah Health operating room locations on or after January 1, 2016 and on or before December 31, 2018. We excluded cases with ASA 5 or 6, age <18 years, cases performed without general anesthesia, as well as obstetric, electroconvulsive therapy, and bronchoscopy procedures. We only included cases where geocoding could match to an address. We limited our study area further to State of Utah counties along the Wasatch Front, as these counties episodically experience some of the worst particulate matter pollution in the US, and globally, due to inversion events where cold air becomes trapped along the mountain valleys, and are also impacted by wildfire smoke in the summer^31,32^. As such, we included only patients residing in the Counties of Salt Lake, Utah, Davis, Weber, Cache, and Box Elder, all of which are served by the University of Utah Health locations in Salt Lake and Davis Counties of Utah. Further exclusions were made based on availability of the PM_2.5_ estimates and ability to assign Elixhauser Comorbidity scores (Figure 2).

**Figure 2:**
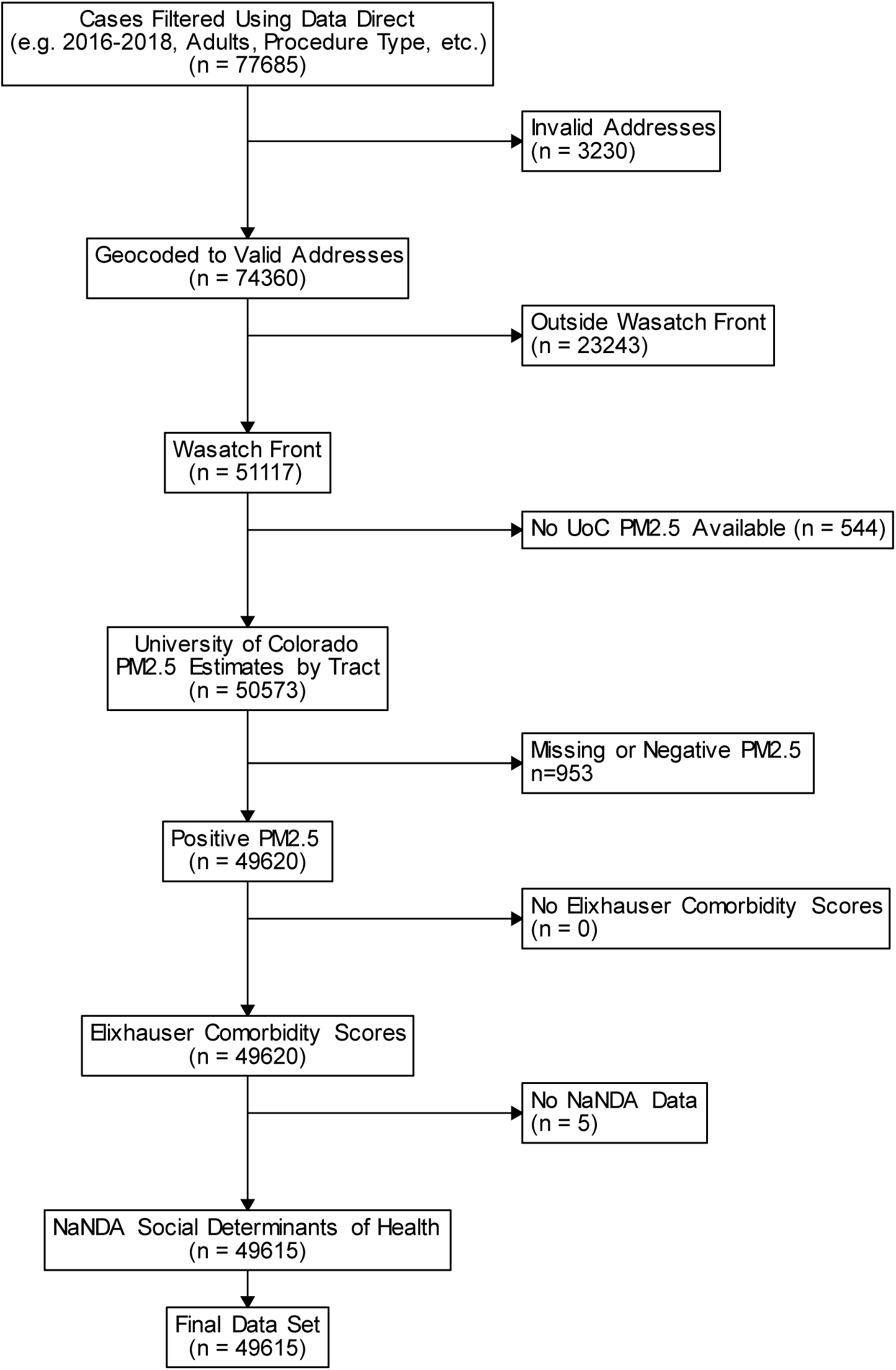
Consort Flow Diagram. This figure details our initisal cohort downloaded from Data Direct from MPOG.org and the process of exclusion criteria applied to generate our final cohort. Of note, the greatest number of patients eliminated was when excluding those who lived outside of the Wasatch Front study area.

### Fine Particulate Matter (PM2.5) Exposure

Daily fine particulate matter with diameter <2.5μm (PM2.5) measurements were obtained from a public dataset of machine learning-derived daily PM_2.5_ concentration estimates at the County, Zip Code, and Census Tract for 11 Western States 2008-2018^33^. These estimates utilize a combination of EPA and state-level ground sensors and satellite derived pollution estimates to provide validated and accurate concentrations of PM_2.5_ across the Western US. We then matched these estimates to individual patients at the census tract of their geocoded home address. For our primary analysis, we utilized the maximum value of PM_2.5_ within 7 days of surgery, which included the day of surgery itself, for two reasons: (1) inversion and pollution events in northern Utah tend to be of short duration^32^, so as to ensure catchment of short-lived events without obscuring them with use of multiple day means, the maximum value in the window was used and (2) prior literature suggests pollution effects on inflammation and thrombosis may manifest over 1-4 weeks, but most acutely within days of the exposure event^34,35^.

### Outcome Bundle: Major Post-operative Complications

The primary outcome was a composite of major postoperative complications occurring during in-hospital stays after surgery as derived from their presence in the discharge diagnosis codes captured in Epic and our local MPOG database^36–38^, including: pneumonia, surgical site infection, urinary tract infection, sepsis, stroke, myocardial infarction, or thromboembolic event. We classified the presence of any post-operative complication into a binary outcome measure, with a positive (yes) if any complication was present, while negative (no) if no complications were observed.

### Covariates

Multivariable models were adjusted for patient age, sex, year, season, County of residence, neighborhood deprivation (which is an index of poverty, race, education and income generated by the National Neighborhood Data Archive (NANDA)^39^), and the Elixhauser comorbidity index^27,28^. Elixhauser comorbidity index was utilized in place of the American Society of Anesthesiologists Physical Classification Score (ASA-PS) as it has demonstrated equivalent reliability in predicting post-operative outcomes^40–42^, and as it is a validated model that relies on documented comorbidities that can be calculated based on Epic data rather than subjective assessments at the bedside. We assigned Elixhauser comorbidity scores to all patients utilizing their EPIC records following standard assignment procedures as outlined by Syed et al^43^. We hypothesized that complications would rise with increasing Elixhauser comorbidity index score.

### Statistical Analysis

We investigated the impact of PM_2.5_ exposure on postoperative complications (outcome) using a multivariable Bayesian model. We utilized a three-season exposure model to adjust for possible confounding factors, where summer months (Jun-Aug) were considered “fire season”, winter months (Dec-Mar) were considered “inversion” season, and other months (Sep-Nov, Apr-May) were considered a baseline period. Thus, all months were included in the model.

Model fit was by hierarchical Bayesian regression methods using Markov chain Monte Carlo algorithms, specifically Hamiltonian Monte Carlo with the No-U-Turn Sampler having more rapid convergence for high-dimensional models. Models were run with six chains, 2000 iterations and a 50% thinning of the initial estimates. For the beta regression parameters, we used two different weakly informative prior distributions to test for sensitivity of parameter estimates. These were the standard normal distribution N(0, 1) and the R2D2M2(0.25, 4, 05) prior^44^. The priors for the variance parameters with the exponential (exp(1)) and the R2D2M2(0.25, 4, 05) priors^44^. County was included in the statistical model as group effect.

Convergence characteristics of the model estimation was assessed using the Gelman-Rubin R-hat statistic, effective sample size (ESS), chain mixing, and chain autocorrelation. The posterior predictive distribution was used to generate a predictive accuracy metric as measured by leave-one-out cross-validation. Nested models were compared by expected-log-predictive-density. Model fits and parameter values were explored using conditional effects, R^2^ coefficient of determination, and Bayesian hypothesis testing.

Model results are presented as parameter estimates and odds ratio transformed values using means, medians, standard deviations, and 95% credible intervals (CI). A 95% credible interval has a 95% probability of containing the true parameter value. Model coefficients are also presented with forest plots to show the probability of direction. Analyses were conducted using R Statistical Software v4.4.1, and used two software packages *built on STAN* (*brms*) and a Hamiltonian MonteCarlo based software to estimate Bayesian models^45^. Additional data and model description was done in the R language using the tableone, loo, mcmcplot, posterior, and tidybayes packages.

## Results

### Study Population Characteristics

Our initial cohort of patients was n=77,685 for the study period of January 1, 2016 to December 31, 2018, as downloaded and filtered from MPOG.org DataDirect. The majority of exclusions occurred from limiting the sample to the Wasatch Front counties (n=23,243), while a small amount of exclusions occurred due to invalid addresses (n=3,230) and missing PM_2.5_ data (n=544), for a final cohort of n=49,615. Median age was 53.22 years (SD=17.67) and a majority were female (52.2%). Most patients had low comorbidity burden, with a mean Elixhauser of 0.89 (SD=1.95). A majority of patients presented from Salt Lake County (n=34,399), which is the most populous county in the state. There was also a slight increase in total cases per year from 15,375 in 2016, to 16,704 in 2018. Table 1 presents baseline characteristics of patients by study year.

**Table 1:** Cohort Characteristics by Year. Overall cohort presented by year. Complications fell marginally year over year, while overall number of cases remained relatively steady. The majority of cases were in Salt Lake County, where the greatest number of University of Utah operating rooms are located.

| Factor | Level | Overall | 2016 | 2017 | 2018 | p-test | SMD |
| --- | --- | --- | --- | --- | --- | --- | --- |
| Sample Total |  | 49615 | 15375<br>(30.9) | 17536<br>(35.3) | 16704<br>(33.6) |  |  |
| Complications (%) | No | 47204<br>(95.1) | 14586<br>(94.9) | 16661<br>(95.0) | 15957<br>(95.5) | 0.014 | 0.02 |
|  | Yes | 2411<br>(4.8) | 789<br>(5.1) | 875<br>(5.0) | 747<br>(4.5) |  |  |
| Elix (mean (SD)) |  | 0.89<br>(1.95) | 0.83<br>(1.83) | 0.91<br>(2.02) | 0.91<br>(1.99) | <0.001 | 0.029 |
| Sex (%) | Male | 23704<br>(47.8) | 7339<br>(47.7) | 8366<br>(47.7) | 7999<br>(47.9) | 0.939 | 0.002 |
|  | Female | 25911<br>(52.2) | 8036<br>(52.3) | 9170<br>(52.3) | 8705<br>(52.1) |  |  |
| Season (%) | None | 17044<br>(34.4) | 4930<br>(32.1) | 5980<br>(34.1) | 6134<br>(36.7) | <0.001 | 0.075 |
|  | Fire | 16472<br>(33.2) | 5457<br>(35.5) | 5704<br>(32.5) | 5311<br>(31.8) |  |  |
|  | Cars | 16099<br>(32.4) | 4988<br>(32.4) | 5852<br>(33.4) | 5259<br>(31.5) |  |  |
| Disadvantage (mean (SD)) |  | 0.06<br>(0.04) | 0.06<br>(0.04) | 0.06<br>(0.04) | 0.06<br>(0.04) | 0.214 | 0.013 |
| Age (mean (SD)) |  | 53.22<br>(17.6) | 53.31<br>(17.61) | 53.43<br>(17.73) | 52.92<br>(17.67) | 0.023 | 0.019 |
| County Name (%) | Box Elder | 672<br>(1.4) | 193<br>(1.3) | 246<br>(1.4) | 233<br>(1.4) | 0.029 | 0.034 |
|  | Cache | 1098<br>(2.2) | 335<br>(2.2) | 386<br>(2.2) | 377<br>(2.3) |  |  |
|  | Davis | 7047<br>(14.2) | 2055<br>(13.4) | 2526<br>(14.4) | 2466<br>(14.8) |  |  |
|  | Salt Lake | 34399<br>(69.3) | 10827<br>(70.4) | 12113<br>(69.1) | 11459<br>(68.6) |  |  |
|  | Utah | 3847<br>(7.8) | 1179<br>(7.7) | 1386<br>(7.9) | 1282<br>(7.7) |  |  |
|  | Weber | 2552<br>(5.1) | 786<br>(5.1) | 879<br>(5.0) | 887<br>(5.3) |  |  |

### Post-Operative Complications

The overall rate of the composite complication outcome was 4.85% (n=2,411 events among total cohort of 49,615); complication rates are presented in Table 2 broken down by population characteristics in a bivariate analysis (yes vs. no complications). The complication rate varied from a low of 4.41% in Davis County to a high of 7.58% in Box Elder County. While female patients dominated the cohort, they were less likely to experience complications than males (3.86% female vs. 5.94% male respectively). As expected, complication rates increased with a greater Elixhauser comorbidity index, with those without complications having a mean index of 0.6 (SD=1.52) while those with complications had a mean index of 5.15 (SD=3.72) (p<0.001). We also observed a slight downtrend in complications over time, from 5.13% in 2016 to 4.47% in 2018 (p=0.014). Complications did not vary significantly by season (p=0.834). Finally, we observed that when PM_2.5_ was dichotomized by the EPA daily limit of 35ug/m^3^, there was an increased complication rate, from 4.8% (n=2332/48468) on days below 35 ug/m^3^ PM_2.5_, to 6.2% (n=79/1147) on days above 35 ug/m^3^ (SMD=0.089), and when this entered into the full model had a posterior distribution of 93.2%. The exposure to high pollution did not differ greatly between Counties, nor did it vary by season. These results are summarized in table 3.

**Table 2:** Cohort characteristics dichotomized by presence of a complication. Cohort dichotomized by presence of complication. The complication rate fell slightly (p=0.014) from 2016 to 2018, with an overall rate of 4.85%. The Elixhauser comorbidity score was markedly higher in for those with complications (5.15) than those without (0.6), while age only varied slightly by those without (53.11) and with (55.28) complications.. Complication rates by County also varied, with Box Elder (7.58%) having the greatest rate and Davis with the lowest (4.41%).

| Factor | Level | Overall | No | Yes | Rate | p-test | SMD |
| --- | --- | --- | --- | --- | --- | --- | --- |
| n |  | 49615 | 47204 | 2411 | 4.85% |  |  |
| Complications = Yes (%) |  | 2411 (4.8) | 0 (0.0) | 2411 (100.0) |  | <0.001 |  |
| Procedure Year Factor (%) | 2016 | 15375 (31.0) | 14586 (30.9) | 789 (32.7) | 5.13% | 0.014 | 0.062 |
|  | 2017 | 17536 (35.3) | 16661 (35.3) | 875 (36.3) | 4.98% |  |  |
|  | 2018 | 16704 (33.7) | 15957 (33.8) | 747 (31.0) | 4.47% |  |  |
| Elix (mean (SD)) |  | 0.89 (1.95) | 0.6 (1.52) | 5.15 (3.72) |  | <0.001 | 1.576 |
| Sex = Female (%) |  | 25911 (52.2) | 24909 (52.8) | 1002 (41.6) | 3.86% | <0.001 | 0.226 |
| Season (%) | None | 17044 (34.4) | 16207 (34.3) | 837 (34.7) | 4.91% | 0.834 | 0.013 |
|  | Fire | 16472 (33.2) | 15667 (33.2) | 805 (33.4) | 4.88% |  |  |
|  | Cars | 16099 (32.4) | 15330 (32.5) | 769 (31.9) | 4.77% |  |  |
| Disadvantage1317 (mean (SD)) |  | 0.06 (0.04) | 0.06 (0.04) | 0.07 (0.04) |  | <0.001 | 0.218 |
| Age (mean (SD)) |  | 53.22 (17.67) | 53.11 (17.72) | 55.28 (16.59) |  | <0.001 | 0.126 |
| County Name (%) | Box Elder | 672 (1.4) | 621 (1.3) | 51 (2.1) | 7.58% | <0.001 | 0.106 |
|  | Cache | 1098 (2.2) | 1041 (2.2) | 57 (2.4) | 5.19% |  |  |
|  | Davis | 7047 (14.2) | 6736 (14.3) | 311 (12.9) | 4.41% |  |  |
|  | Salt Lake | 34399 (69.3) | 32775 (69.4) | 1624 (67.4) | 4.72% |  |  |
|  | Utah | 3847 (7.8) | 3612 (7.7) | 235 (9.7) | 6.10% |  |  |
|  | Weber | 2552 (5.1) | 2419 (5.1) | 133 (5.5) | 5.21% |  |  |

**Table 3:** Cohort characteristics dichotomized by whether or not exposed to maximum of 35ug/m^3^ in pre-operative period. Table 3 dichotomizes patients based on their exposure to any day of PM_2.5_ greater than 35ug/m^3^ in the 7 day pre-operative period. In this instance, the complication rate was higher when patients were exposed to PM_2.5_>35 (SMD=0.089). Notably, 2017 had much higher exposure to elevated PM_2.5_ (546 Cases, 47.6% of total exposures), while disadvantage was not dramatically different between higher (0.07) and lower (0.06). Finally, exposure to fire vs. cars as source of pollution was nearly evenly split, indicating equal exposure to elevated inversion pollution and wildfire smoke.

| Factor | Level | Overall | Max PM2.5<br>> 35 | Max PM2.5<br><35 | SMD |
| --- | --- | --- | --- | --- | --- |
| n |  | 49615 | 1147 | 48468 |  |
| Complications (%) | No | 47204 (95.1) | 1068 (93.1) | 46136 (95.2) | 0.089 |
|  | Yes | 2411 (4.9) | 79 (6.9) | 2332 (4.8) |  |
| Procedure Year Factor (%) | 2016 | 15375 (31.0) | 274 (23.9) | 15101 (31.2) | 0.260 |
|  | 2017 | 17536 (35.3) | 546 (47.6) | 16990 (35.1) |  |
|  | 2018 | 16704 (33.7) | 327 (28.5) | 16377 (33.8) |  |
| Elix (mean (SD)) |  | 0.89 (1.95) | 1.02 (2.00) | 0.88 (1.95) | 0.067 |
| Sex (%) | Male | 23704 (47.8) | 549 (47.9) | 23155 (47.8) | 0.002 |
|  | Female | 25911 (52.2) | 598 (52.1) | 25313 (52.2) |  |
| Season (%) | None | 17044 (34.4) | 0 (0.0) | 17044 (35.2) | 1.042 |
|  | Fire | 16472 (33.2) | 578 (50.4) | 15894 (32.8) |  |
|  | Cars | 16099 (32.4) | 569 (49.6) | 15530 (32.0) |  |
| Disadvantage (mean (SD)) |  | 0.06 (0.04) | 0.07 (0.04) | 0.06 (0.04) | 0.310 |
| Age (mean (SD)) |  | 53.22 (17.67) | 52.26 (17.80) | 53.24 (17.67) | 0.056 |
| County Name (%) | Box Elder | 672 (1.4) | 18 (1.6) | 654 (1.3) | 0.058 |
|  | Cache | 1098 (2.2) | 29 (2.5) | 1069 (2.2) |  |
|  | Davis | 7047 (14.2) | 175 (15.3) | 6872 (14.2) |  |
|  | Salt Lake | 34399 (69.3) | 786 (68.5) | 33613 (69.4) |  |
|  | Utah | 3847 (7.8) | 77 (6.7) | 3770 (7.8) |  |
|  | Weber | 2552 (5.1) | 62 (5.4) | 2490 (5.1) |  |

### Multivariable Exposure Analysis of PM_2.5_ impact on outcome bundle

Model estimation satisfied usual criteria (Gelman-Rubin R-hat, ESS, posterior predictive error checks). There was no significant autocorrelation. The posterior density plots for model parameters indicate reasonable unimodal distributions. Our sensitivity analysis exploring both two weakly informative priors did not change the inferences or results. Our findings were robust to model parameters and model specifications.

In our main Bayesian multivariable model, we found an increased risk of post-operative complications with increasing concentrations of PM_2.5_ with a regression coefficient estimate of 0.01 with a 95% CI (0.00-0.01). A Bayesian hypothesis test showed a 99% probability of an increasing probability of complications with increasing PM_2.5_. The Odds Ratio(OR) of a major perioperative complication for a one unit increase in PM_2.5_ is 1.0079, and for a ten unit increase the OR is 1.082. In clinical terms there is an 8% increase in the chance of complication for every 10ug/m^3^ increase in the maximum PM2.5 observed in the 7-day preoperative period. For a 30 fold increase in PM_2.5_ (1 ug/m^3^ to 30ug/m^3^) the odds of complication is over 27% (95%CI: 4%-55%). This increase was noted in a curvilinear fashion, as presented in Figure 3. There was no apparent change in this relationship across seasons as we defined them for this region (fire, inversion, baseline), nor did the inclusion of neighborhood disadvantage alter the findings. The impact of PM_2.5_ was of greater magnitude among patients with a higher Elixhauser comorbidity score, especially for those with Elixhauser of 3 or greater compared to those with Elixhauser <1, as shown in Figure 3. Overall the parameters in the model explained about 1/4^th^ of the variance (R2 = 0.28).

**Figure 3:**
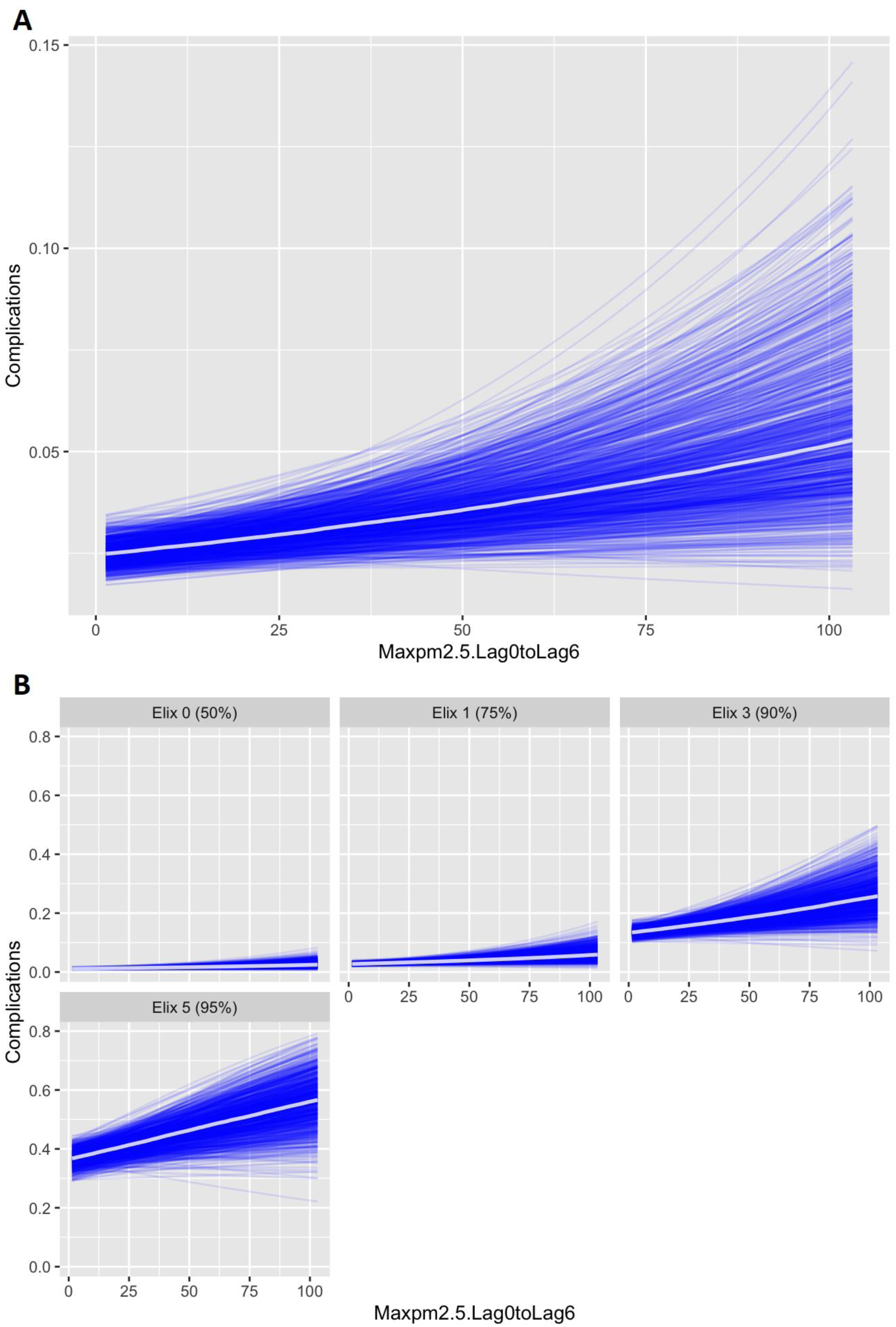
Maximum PM2.5 in the 7 days pre-operatively (Lag0-Lag6) vs. Complication Rate. Panel A (top) details Complications Rate vs. Maximum PM2.5 of the 7-day lag period, while Panel B (bottom) details the same broken down by Elixhuaser Comorbidity Index. Complication rate is shown as absolute value, so here 0.04 = 4% complication rate. The PM2.5 values are the maximum observed in the 7-day preop window, in ug/m^3^, Lag 0 is day of surgery. In Panel B as noted, the overall complication rate for those with higher Elixhauser comorbidity index is higher at baseline, but rose more quickly with elevations in PM2.5. This response to PM2.5 continued to increase in magnitude as Elixhauser index increased. Note that the effect of PM2.5 was greater among this with Elixhauser of 5, but that the baseline complication rate was also higher.

## Discussion

### Summary of Findings

In our single center cohort of over 49,000 patients undergoing elective surgery at an academic medical center near the Wasatch front in Utah, we found that increased exposure to PM_2.5_ in the 7-days prior to surgery was associated with significantly higher risk of postoperative complications, confirming our primary hypothesis. The findings were statistically highly significant, and the effect size is consistent with risk observed in the air pollution epidemiology literature^46^, providing further confirmation of our findings. We observed a dose-response relationship, with over 27% increased risk of complications at the highest PM_2.5_ concentrations compared to low concentrations, with significant increase in risk above an apparent cut-off exposure of 35 ug/m^3^ (Table 3). This association and the cutoff were consistent across all patient age groups, with individuals with higher Elixauser Co-morbidity status appearing to have greater susceptibility to elevated PM_2.5_ (Figure 3). Age, sex, County, year of procedure, neighborhood deprivation (a measure of social distress) and season appeared to have minimal influence on the association. We found that the rate of our complication bundle decreased year over year, while the distribution of Elixhauser scores remained consistent, and the exposure to air pollution remained comparable between years.

### Implications for clinical practice, research and policy

To our knowledge, this is the first large-scale study demonstrating the acute impact of preoperative IPAPE on postsurgical inpatient outcomes in a large surgical patient cohort in an OECD country^22,26^. The implications are threefold; our study should inform: (1) Policy, demonstrating a potentially novel and unaccounted for susceptible population (surgery patients), (2) Individual patient scheduling decisions for high-risk patients (elevated Elixhauser index) and targeted mitigation (masks, filters), and (3) Further research into the pathophysiological mechanisms of healing after surgery in human and animal studies.

### Policy Implications

While air pollution in some regions of the U.S. have improved dramatically since the 1970s, improvements have often benefitted certain groups over others. For instance, Black Americans are exposed to higher annual concentrations of air pollution containing fine particulate matter than their White counter parts^47^. Minority populations often face greater pollution burdens and may also be more susceptible to their health effects due to their lack of access to health services and lower income and educational status, which reduces accessibility to pollution information^47^ and other forms of prevention. This problem tracks globally as well. Today, air pollution is considered a leading causes of health complications and mortality worldwide, especially affecting lower-income groups, who tend to be more exposed and vulnerable^48^. Consequently, the findings presented in this research have important policy implications for public health and contribute to the literature on health disparities.

Climate change is expected to increase the incidence of wildfires by 29% worldwide^49^, which will disproportionately burdening urban and developing regions. These regions are already facing a growing severity of pollution impacts; this research suggests that the interaction of surgery and pollution will also lead to increasing health burdens. For instance, in China where approximately 10% of GDP^50^ is spent on the health effects of pollution, a 1% increase of PM_2.5_ currently leads to a 2.942% increase in household healthcare expenditure^51^ and these numbers are expected to grow over the next ten years. Subsequently, developing states, global megacities, and urban areas in Asia are already faced with tough and costly choices. Economically, the impact of air pollution on surgery outcomes will further contribute to decreases in labor productivity for patients and their families, and, therefore, a lower tax base for these communities. In addition, poor surgical outcomes lead to inefficient use of hospital resources, increases the costs of insurance for all, and maybe lead to long term welfare costs, especially where loss of life is concerned. When combined with the disproportionate health disparities, the scale of costs are dramatic for all major population areas.

### Scheduling and Targeted Mitigation

Targeted mitigation efforts around the perioperative period may reduce complications attributable to PM_2.5_ exposure among susceptible patients. Surgical delay or protection from pollution during high-risk periods through use of indoor HEPA filtration systems could be considered for patients with planned procedures (if they work indoors or from home) during seasonal inversion events or wildfire smoke conditions. Huang et al’s study from China^23^, though it focused on mortality, found a beneficial economic impact from rescheduling elective surgery cases in at-risk populations during high pollution events. The implications for elective and non-urgent major surgery, especially during wildfire events, merit further investigation with multidisciplinary teams.

### Research Implications and Future Directions

Our findings lead to several new research questions. Our ability to identify high risk populations through sub-group analyses was limited due to sample size, and extension of our study to more years and more sites should help elucidate questions regarding the most vulnerable surgical populations. Additional outcomes should be studied to corroborate our findings and refine the impact of air pollution, incorporating outcome measures such as length of stay, mortality, and additional long- and short-term outcomes. Ours and others future epidemiological studies should also guide and be balanced with animal models where more precise biochemical mechanisms can be uncovered. We are also concerned that social determinants of health might confound the association of small particle pollution with post-surgical outcomes and will study this association in subsequent investigations. Finally, while we did identify that the relationship with PM_2.5_ appeared to accelerate with higher concentrations of pollution, formulating this exposure mathematically into a functional relationship will be a challenge for future research.

### Strength and Limitations

This study has several strengths:

1. The natural geography of Utah creates a unique strength to this study in that the same population is exposed to low and high concentrations of pollution.
2. We examined a large cohort of over 49,000 patients over a 3-year contemporary period. We utilized precise PM_2.5_ estimates at the small census tract level and investigated the effects across the full range of PM2_2.5_ concentrations. We tested different exposure windows and adjusted for clinical comorbidities.
3. Our outcome measure was a previously used and validated composite of serious complications encompassing major morbidity events. We specifically focused on major morbidity complications that are meaningful outcomes for quality improvement and risk mitigation efforts around elective surgery timing as well as patient optimization and that are plausibly related to IPAPE. We controlled for individual patient comorbidities with the validated and widely used Elixhauser comorbidity index.

Our study has some limitations:

1. Exposure: While our study leveraged a large sample size, we estimated IPAPE based on census tract locations. This model does not account for proximity to highways, industrial sources, or other sources of PM_2.5_ pollution that may be more chronic rather than episodic in nature. This model also used daily mean exposure estimates and lacked elevation in the model. Both factors could be biased in terms of missing peaks of exposure within 24-hour windows and at different elevations, which could lead to further bias in exposure estimates. Additionally, we evaluated PM_2.5_ mass concentration and did not have data on particulate composition that may influence toxicity^52^.
2. Mitigation and social determinants of health: We could not control for in-home filtration or other personal mitigation measures, which may be less available to those of lower incomes, non-English Language speakers, and in more socially vulnerable neighborhoods.
3. Population: Our cohort was predominantly white and treated at a single health system, although population-level variability in PM_2.5_ exposure was leveraged. We did not account for patient reported or EHR recorded race and ethnicity, which may confound our results. Despite the Salt Lake City region’s reputation for homogeneity, the metropolitan area is close to the median diversity index for mid-size US cities. As a result, the findings of this study could be generalized to similar metropolitan areas, both larger and smaller, though the region is dominated by white and Hispanic populations.
4. Analysis Type: As a retrospective analysis, unmeasured confounding is also a possibility, as is incorrect inferences due to the ecological fallacy, we discussed in detail elsewhere^53^, though the use of residential address mitigates this in part.

### Comparison with the literature and proposed mechanistic pathways

Our findings from over 49,000 patients along Utah’s heavily polluted Wasatch Front build on limited prior data on air pollution and postoperative outcomes. In the China analysis, city-level PM_2.5_ during the week prior to surgery was associated with 1% higher adjusted 30-day mortality per 10 μg/m^3^ increase in PM_2.5_^23^. Their mortality association was stronger at higher PM_2.5_ levels (>100 μg/m^3^), similar to our findings. The few other studies that have examined adverse impacts of air pollution on perioperative outcomes have been limited in scope, methodologies and patient populations: A majority of these studies examined only organ transplants^26^: while a reasonable suspicion given their immunocompromised nature, they are a relatively small and high-risk patient populations thus limiting generalizability. Spencer-Hwang et al found that kidney transplants have an increased risk of fatal MI with increasing ozone, in a dose-dependent manner^54^, while studies of lung transplant recipients suggest that proximity to major roads can increase risk of chronic allograft dysfunction^26^. Recent data from California also suggests pediatric patients are susceptible to adverse pulmonary events under anesthesia during wildfire events, especially those with reactive airway disease^25^.

The 7-day PM_2.5_ exposure timeframe corresponds to the period when detrimental effects of pollution on inflammatory, thrombotic, and immune pathways implicated in surgical complications may become manifest^55^. For example, PM_2.5_ instigates systemic inflammation through release of IL-1, IL-6, and CRP as well as reactive oxygen species^14,56^. Resultant endothelial dysfunction promotes a prothrombotic state over 7-14 days^57^. Surgical trauma induces a similar acute phase response and immunomodulation^10–12^, which combined with the biological impact from recent pollution exposure could synergistically heighten complication risk, as detailed in Appendix Figure 1. Taken as a whole, the literature suggests adverse impacts from air pollution, and most notably fine particulate matter, on outcomes after surgery. Though major gaps exist on which pollution source may be most harmful and which patient population may be most affected. Further studies should explore possible mitigation strategies, either pre-operatively or post-operatively, for example the offering of patient bedroom air filters or patient masks preoperatively, or the rescheduling of elective high-risk patients during extreme exposure events, like wildfires.

## Conclusion

In our single center cohort study, we found that elevated individual patient exposure to small particles in the week prior to surgery was associated with significantly increased postoperative complications. We demonstrated a dose-response relationship in all age groups, regardless of patient co-morbidity (as measured by the Elixhauser index) in a large cohort at an academic medical center in Utah. This clearly demonstrates a statistically significant impact of air pollution exposure on surgical outcomes, with an effect size consistent with the broader pollution literature^46^.

Our results cover a wide swath of surgical specialties and thus have implications for much of elective surgery performed in areas suffering from periodic acute pollution episodes, such as inversion events and, much more commonly with climate change, wildfire smoke. At-risk patients may benefit from pollution mitigation and close monitoring in the postoperative period after procedures preceded by heavy pollution exposure. Overall, our findings highlight that limiting particulate matter exposure through clean air policies and practices can have wide-ranging health benefits beyond just cardiopulmonary disease, including reducing complications of surgery.

## Data Availability

All data produced in the present study are available upon reasonable request to the authors.

## Author Institutional Titles

John F. Pearson MD, Associate Professor of Anesthesiology

Cameron K. Jacobson, MS, Associate Director of Information Technology

Calvin S. Riss, Graduate Research Assistant

Matthew J. Strickland, Professor and Chair of Public Health

Longyin Lee, Graduate Research Assistant

Neng Wan, Associate Professor

Tabitha M. Benney, Associate Professor of Political Science

Nathan L. Pace, Professor of Anesthesiology

Ben K. Goodrich, Associate Research Scholar

Jonah S. Gabry, Senior Research Associate

John V. Pham, Assistant Professor of Anesthesiology

Cade K. Kartchner, Medical Student Year 2 (MS-2)

Jake Wood, Medical Student Year 4 (MS-4)

Michael H. Andreae, Professor of Anesthesiology

## Prior Presentations

Association of Air Pollution with Perioperative Complications, 18^th^ Annual World Congress of Anaesthesiologists, March 5, 2024, Singapore.

## Acknowledgements

The authors would like to acknowledge TELLUS Networked Sensor Solutions, Inc. for the preparation and provision of the data and visualizations in Figure 1.

## Funding Statement

This study was funded from an intramural University of Utah Wilkes Center for Climate Science and Policy seed grant [to JFP and NW] as well as partially supported by the National Institute of Health National Institute of Environmental Health Sciences and National Cancer Institute (NIH grant 5R01ES029528-05 [to MS], 5R37CA276365-02 [to NW]). Further funding was from the National Science Foundation (NSF grant 2051246 [BG] and 2153019[BG]). The funding sources were not involved in the study design, collection, analysis and interpretation of the data, in writing the report, or in the decision to submit the article for publication. The content is solely the responsibility of the authors and does not necessarily represent the official views of the National Institutes of Health, the National Science Foundation, or the University of Utah Wilkes Center for Climate Science and Policy.

## Conflicts of Interest

Ben Goodrich and Jonah Gabry are both member-managers of GG Statistics, LLC. The remainder of the authors declare no competing interests.

## Use of Artificial Intelligence Tools

During the preparation of this work the authors used OpenAI’s ChatGPT for assistance with writing clarity and literature search. After using this service, the authors reviewed and edited the content and take full responsibility for the content of this publication. This information is included in the Methods section of the manuscript.

## Author Approval

All authors have seen and approved the manuscript.

## Notes

### Author Declarations

The Institutional Review Board of the University of Utah gave ethical approval of this work as Category 4, Exempt with IRB approval number 00142167.

### Summary of Updates

Revision for submission to new journal, and update to the cohort size after discovery of data wrangling issues. All analysis were re-run with new cohort, and results found to be highly similar (in magnitude and direction) and conclusions essentially unchanged from prior posting.

