## Appendix Figure 1 for "Preoperative Exposure to Fine Particulate Matter and Risk of Postoperative Complications: A Single Center Observational Cohort Bayesian Analysis"

**Supplemental Figure 1 (Breakout Box): Mechanism interaction between air pollution exposure and systemic inflammatory response.**

**Legend:**

The components of fine particulate matter ($PM_{2.5}$) and their insidious impacts on organ systems, as well as their synergy with the surgical stress response (SRS), are detailed in this diagram. $PM_{2.5}$ is show at the top, with the dominant sources in triangles (vehicles, industrial facilities, furnaces, terrestrial sources, and fossil fuel power), along with the various components (Potassium, Copper, etc.). SRS results in sympathetic activation, increasing heart rate and blood pressure (noted in the cardiac box), while reducing blood flow to the kidneys (noted in the renal box)[10,11,13]. These responses increase the risk of stroke through coagulation (noted in the neurological box)[12,14], as well as myocardial infarction and acute kidney injury. Kidneys are further inflamed by release of TNF-alpha and direct oxidative stress damage from pollutants[16]. The lungs are most directly impacted by pollutants, with a direct inflammatory response at the alveoli, which can synergize with the barotrauma and volutrauma inflammation commonly seen from positive pressure ventilation. In the immune system (bottom right panel) there is emerging evidence that particulates exacerbate the T-helper 2 responses, which is also seen in later phases of the SRS, thus the potential increased risk for infectious complications[10,11,14,57].

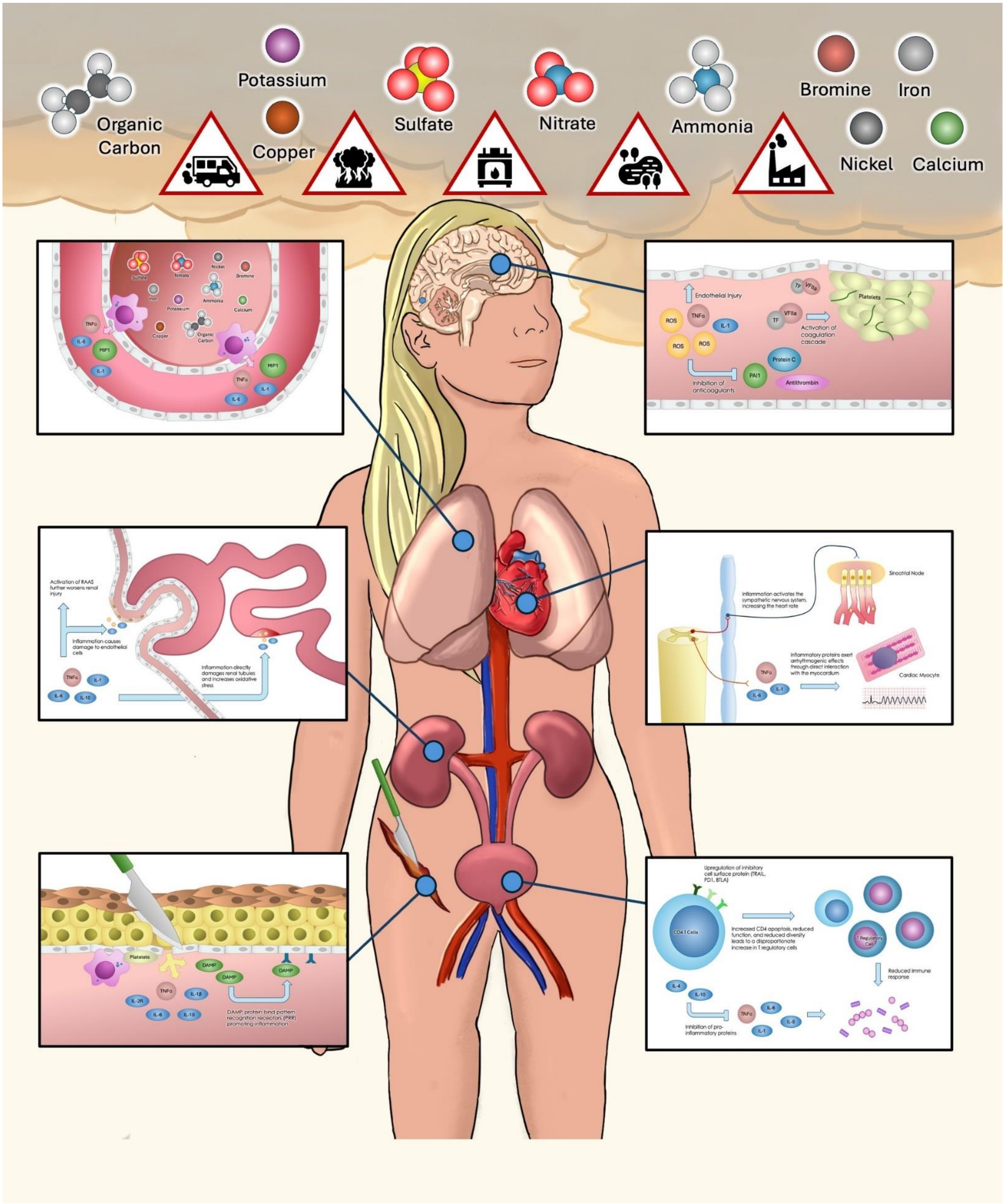

Organic Carbon
Potassium
Copper
Sulfate
Nitrate
Ammonia
Bromine
Iron
Nickel
Calcium
Endothelial Injury
TNFα
ROS
IL-1
TF
VFIIa
Platelets
Activation of coagulation cascade
Protein C
PAI1
Antithrombin
Inhibition of anticoagulants
MIP1
IL-6
Activation of RAAS further worsens renal injury
Inflammation causes damage to endothelial cells
Inflammation directly damages renal tubules and increases oxidative stress
IL-10
Inflammation activates the sympathetic nervous system, increasing the heart rate
Sinoatrial Node
Inflammatory proteins exert arrhythmogenic effects through direct interaction with the myocardium
Cardiac Myocyte
DAMP
IL-1β
IL-2R
DAMP protein bind pattern recognition receptors (PRR) promoting inflammation
Upregulation of inhibitory cell surface protein (TRAIL, PD1, BTLA)
CD4 T Cells
Increased CD4 apoptosis, reduced function, and reduced diversity leads to a disproportionate increase in T regulatory cells
T Regulatory Cell
Reduced immune response
IL-4
Inhibition of pro-inflammatory proteins
