## Appendix Table 1 for "Preoperative Exposure to Fine Particulate Matter and Risk of Postoperative Complications: A Single Center Observational Cohort Bayesian Analysis"

| Author / Publication Year | Population | N | Exposure | Comparison | Outcome | Study Design | DOI |
|---|---|---|---|---|---|---|---|
| **United States** | | | | | | | |
| Chang SH. 2021 | Post Renal Transplant | 112,098 | Annual PM2.5 | Annual mean PM2.5 Gradient | Acute Rejection, Mortality | Retrospective Cohort | 10.1001/jamanetworkopen.2021.28568 |
| Dehom. 2021 | Post Renal Transplant | 93,857 | Annual PM2.5 | Race (Black vs. Non-Black RTR) | Mortality, CVE, Mortality from CHD | Retrospective | 10.3390/ijerph18084297 |
| Feng, Y. 2021 | Post Kidney Transplant | 87,223 | Annual PM2.5 | Exposure by zip code | DGF, Acute Rejection, All-cause mortality | Retrospective | 10.1111/ajt.16605 |
| Spencer-Hwang. 2011 | Non-Smokers Post Renal Transplant | 38,101 | Annual Ozone, PM10 | Gender, Exposure by zip code | Mortality from CHD | Retrospective Cohort | 10.4172/2155-9880.S6-001 |
| Spencer-Hwang. 2011 | Non-Smokers Post Renal Transplant | 32,239 | Annual Ozone, PM10 | Concentrations by zip code | CHD Death, mortality | Retrospective Cohort | 10.1053/j.ajkd.2011.05.017 |
| Al-Kindi SG. 2019 | Post Heart Transplant | 21,800 | Annual PM2.5 | Exposure gradient | Mortality | Retrospective Cohort | 10.1016/j.jacc.2019.09.066 |
| Marsh BJ. 2022 | Pediatric patients | 625 | **AQI>100 in 2 weeks prior to surgery** | AQI<100 in 2 weeks prior to surgery | Adverse Respiratory events | Retrospective Double-Cohort | 10.1097/aln.0000000000004344 |
| **Canada** | | | | | | | |
| Bhinder S. 2014 | Post Lung Transplant | 397 | Major Roadway Proximity, Annual Ozone, PM2.5, NO2 | Major Roadway Proximity, Annual Concentrations Gradient | CLAD, Mortality | Retrospective Cohort | 10.1111/ajt.12909 |
| **Europe** | | | | | | | |
| Pierotti, L. 2018 | Post Solid Organ Transplant | 13,959 | Major Roadway Proximity, Annual: PM2.5, PM10, NO2, NOx | Major Roadway Proximity, Annual Concentration Gradient | Mortality, Renal Graft Failure | Retrospective Cohort | 10.1016/j.jth.2018.05.100 |
| Ruttens D. 2017 | Post Lung Transplant | 5,707 | Annual PM10, Major Roadway Proximity | Major Roadway Proximity, PM10 exposure at residential address | CLAD, Mortality | Retrospective Cohort | 10.1183/13993003.00484-2016 |
| Verleden SE. 2012 | Post Lung Transplant | 1,276 | **7-day lag of daily PM10 by residential address** | Azithromycin vs Non-Azithromycin; Pollution Gradient | A-Grade Rejection, LB and BAL | Prospective observational | 10.1111/j.1600-6143.2012.04134.x |
| Benmerad M. 2017 | Post Lung Transplant | 520 | Annual PM2.5, PM10, Ozone, NO2 | Azithromycin vs Non-Azithromycin; Pollution Gradient | FEV1 | Prospective longitudinal | 10.1183/13993003.00206-2016 |
| Nawrot TS. 2011 | Post Lung Transplant | 288 | Annual PM10, Major roadway proximity | Annual PM10, Major Roadway Proximity | BOS, Mortality | Retrospective Cohort | 10.1136/thx.2010.155192 |
| **Asia** | | | | | | | |
| Huang J. 2023 | Surgical patients | 1,381,283 | **Acute PM2.5** | Exposure on day of surgery | Mortality, SSI, Cardiopulmonary Complications | Retrospective | 10.1016/j.jpubeco.2023.104825 |
| Oh, TK. 2023 | Patients Post Major Cancer Surgery | 244,766 | Annual PM2.5, PM10, Ozone, NO2, SO2, CO | Gradients of Annual Exposure at home address | 90-Day, 1-Year, and All-Cause Mortality | Retrospective Cohort | 10.1097/jom.0000000000003009 |
| Park, JB. 2024 | Children Receiving General Anesthesia | 13,175 | **Acute PM2.5, PM10**, | Gradients of Exposure on day of surgery | Intra-operative hypoxemia | Single Center Retrospective | 10.1097/eja.0000000000002027 |
| Liu, C. 2023 | Lung Cancer Post Lobectomy | 3,327 | Monthly PM2.5, Ozone | Exposure by residential address | Survival | Prospective | 10.1186/s12940-023-00976-x |
| Che, Lu. 2017 | Surgical patients with delirium | 559 | PM2.5, PM10, SO2, NO2, CO | Exposure on day of surgery | Delirium incidence | Time-stratified case-crossover | 10.1038/s41598-017-15280-1 |

### Supplemental Table 1: Rigor of Prior Research.

Less than 20 prior studies investigate the association between air pollution and adverse surgical outcomes, and many were small and limited to specialized populations like organ transplant recipients, surgical oncology patients, and pediatrics[22,26]. A recent analysis of over 1.3 million patients in China found increased 30-day postoperative mortality associated with higher preoperative $PM_{2.5}$ concentrations at the city-level, especially among patients with preexisting cardiopulmonary conditions and those undergoing surgical oncology procedures[23]. Likewise, a recent study from South Korea found a similar mortality risk among cancer patients[24]. However, both of these studies focus on mortality, not morbidity, and pollution levels in China far exceed those encountered in the U.S. or along the Wasatch Front in Utah, where this study took place. A notable difference with our approach is the majority of studies utilize annual or monthly exposures to air pollution, with only four studies utilizing a lag period or acute exposures on day of surgery.

*Abbreviations*: PM2.5, Particulate matter 2.5; PM10, Particulate matter 10; NO2, Nitrogen Dioxide; AQI, Air Quality Index; CHD, Chronic Heart Disease; US, United States; SO2, Sulfur Dioxide; CO, Carbon Monoxide; LB, Lymphocytic Bronchiolitis; BAL, Bronchoalveolar lavage; CLAD, Chronic Lung Allograft Dysfunction; FEV1, Forced Expiratory Volume in one second; CVD, Cardiovascular Disease. RTR, Renal Transplant Recipients.
